# Influenza and other respiratory viral infections associated with absence from school among schoolchildren in Pittsburgh, Pennsylvania, USA: a cohort study

**DOI:** 10.1101/2020.04.16.20068593

**Authors:** Jonathan M. Read, Shanta Zimmer, Charles Vukotich, Mary Lou Schweizer, David Galloway, Carrie Lingle, Gaby Yearwood, Patti Calderone, Eva Noble, Talia Quadelacy, Kyra Grantz, Charles Rinaldo, Hongjiang Gao, Jeanette Rainey, Amra Uzicanin, Derek A.T. Cummings

## Abstract

**Background:** Information on the etiology and age-specific burden of respiratory viral infections among school-aged children remains limited.

**Methods:** We conducted a cohort study to determine the etiology of ILI (influenza like illness) among 2,519 K–12 students during the 2012–13 influenza season. We obtained nasal swabs from students with ILI-related absences. Generalized linear mixed-effect regressions determined associations of outcomes, including ILI and laboratory-confirmed respiratory virus infection, with school grade and other covariates.

**Results:** Overall, 459 swabs were obtained from 552 ILI–related absences. Respiratory viruses were found in 292 (63.6%) samples. Influenza was found in 189 (41.2%) samples. with influenza B found in 134 (70.9%). Rates of influenza B were significantly higher in grades 1 (10.1%, 95% CI 6.8%–14.4%), 2 (9.7%, 6.6%–13.6%), 3 (9.3%, 6.3%–13.2%), and 4 (9.9%, 6.8%–13.8%) than in kindergarteners (3.2%, 1.5%–6.0%). After accounting for grade, sex and self-reported vaccination status, influenza B infection risk was lower among kindergarteners in half-day programs compared to kindergarteners in full-day programs (OR = 0.19; 95% CI 0.08–0.45).

**Conclusions:** ILI and influenza infection is concentrated in younger schoolchildren. Reduced infection by respiratory viruses is associated with a truncated school day for kindergarteners, but requires further investigation in other grades and populations.

## Background

The 2009 influenza pandemic caused disruption to schools, businesses, and governmental entities. Numerous reports from US CDC and European countries document the central role of school-age children in community-wide transmission of pandemic virus [1-10]. In the United States and Australia, the onset of pandemic influenza incidence was linked to school opening dates [11]. Children experience higher rates of infection, shed influenza virus longer than adults, and have social mixing patterns conducive to the propagation of respiratory viruses [5, 12]. However, the risk distribution of influenza and other respiratory infections throughout childhood is poorly understood. Current influenza vaccination programs in the United States target individuals 6 months of age and older. If vaccine supply is limited, children 6-59 months of age are an age group of focus for vaccination [13]. Programs in the United Kingdom target children 17 years of age and younger, with a program that phases universal coverage in beginning with younger children [14].

A number of seasonal pathogens cause respiratory symptoms consistent with influenza-like illness (ILI) [15]. Given the importance of school-aged children for influenza transmission, a better understanding of the etiology of respiratory infections among children, their associated burden, and the interaction of these infections with influenza is required to improve current public health strategies to reduce transmission of infections in schools and communities.

Despite the role of school age children in driving influenza epidemics, relatively little fine-grained analysis has been conducted to assess how different ages within the school demographic are affected by seasonal influenza [16]. Factors other than age may also be important in determining risk of infection. Females have been shown to be at greater risk of influenza infection in adults [17], but it is unknown if school age females are also at elevated risk compared to male pupils in the same schools. Small-scale spatial variation in influenza attack rate has been observed [18], though it is unclear whether nearby schools (within the same district) experience similar epidemic dynamics. The acquisition of immunity during childhood is thought to drive patterns of age specific incidence of influenza [19]; few detailed studies have examined how infection risk changes with age during childhood.

We conducted the SMART study (Social Mixing and Respiratory Transmission) in two school districts in the Pittsburgh metropolitan area. This study included testing school-aged children for respiratory viruses following an ILI-related absence during the winter influenza season of 2012-2013 in a cohort of schools. We relate infection outcomes to attributes of individual children and grade and classroom properties, as well as school districts and the individual schools within them.

## Methods

### Study population

We conducted our study in a charter school system in an urban area (district A: eight schools and 2,000 kindergarten to 12^th^ grade [K-12] students) and a public school district in a suburban area surrounding an urban core (district B: nine schools and approximately 4,700 K-12 students). We worked with nine schools selected from these two districts: three from district A and six from district B.

### Ethical Review

Protocol and all study materials were approved by the University of Pittsburgh IRB #PRO11020500, the University of Florida IRB #IRB201701941, Centers for Disease Control and Prevention IRB #IRB00000319, the University of Liverpool Research Ethics Committee, and the Research Ethics Committee of the Faculty of Health and Medicine at Lancaster University.

### Recruitment and study procedures

Project staff met with schoolboards, parent-teacher organizations, school staff, and school nurses to provide information about the project. Investigators provided students and parents a summary of the study, including a disclosure form and a signature section for opting out of participation. All parents received information sheets ahead of the study period, offering them the option to withdraw their child from the study. All participating children gave verbal assent prior to swabbing and interviewing.

### Surveillance and virologic sampling

The 2012-13 influenza transmission season started unusually early nationwide [20]. In schools participating in our study, influenza cases began to rise in mid-December. The SMART study team initiated surveillance in three schools during the week of December 17-21, 2012, prior to the scheduled winter break, and in the first week of January 2013. Surveillance activities were implemented in all nine participating schools from the start of the spring semester on January 7, 2013, concluding on March 27, 2013. Schools closed for winter break December 24, 2012 to January 2, 2013; spring break closure was March 28 to April 1, 2013.

Trained project staff were deployed in participating schools to monitor student absentee data, receiving daily attendance reports from schools. We defined an absence event as absence from school by a student for either an entire day or missing part of the day due to leaving school early due to illness. This excluded individuals who were tardy but present for part of the day. All contiguous days for which the student was absent were classified as a single absence event. Study staff telephoned parents to determine the reason for student absences. If an absent student was reported ill, staff then inquired about the symptoms the student had (categories recorded were: fever, sore throat, cough, runny nose/congestion, headache, muscle or joint pain, nausea/diarrhea/vomiting). Students with ILI using the CDC Case Definition − fever of at least 37.8 °C, and either cough or sore throat − were eligible for nasal swabbing by project or school staff upon return to school. Temperature was requested and recorded for those individuals who reported a recorded temperature, however, to increase sensitivity of case finding, children that a parent or guardian reported as having a fever, even in the absence of a measurement were included as meeting our case definition. Students presenting to the school nurse with ILI were swabbed immediately, otherwise students were swabbed on their return to school following illness absence.

### Sampling and laboratory analysis

Staff used sterile wound polyester-tip swabs to collect specimens from the anterior nares of participants; the swab was placed in sterile transport media, and transported to University of Pittsburgh Medical Center Clinical Virology Laboratory, where PCR tests were performed for both influenza A and B, and a range of other respiratory viruses (see supplement for further details).

### Questionnaire

Participating students were surveyed on a repeated basis to determine influenza vaccine status.

### Statistical Analysis

Cumulative attack rates (CARs) were calculated from pooled data across all schools and the entirety of the surveillance period. CARs were calculated for participants who were identified as: 1) having ILI, 2) testing positive for any of the respiratory viruses in the panel, or 3) testing positive for influenza virus, regardless of subtype and testing positive for each of subtypes of influenza separately. Denominator values included the number of participating students, students with absence periods who reported to the school nurse, and students with absence periods who did not report to the school nurse but whose home we were able to contact. We excluded students whose homes we were not able to contact to determine their health status even after multiple attempts. Binomial confidence intervals were calculated for all CARs.

We performed a variety of generalized linear mixed-effect regressions, in which outcome variables were the binary infection status of a participant over the entire surveillance period given a particular definition of outcome: whether they were identified as having ILI; testing positive for any respiratory virus; testing positive for influenza; or testing positive for a specific subtype of influenza (A and B). We included hierarchical random intercept terms for school and district in all models unless specified otherwise, where schools were nested within one of the two districts. We performed hypothesis-driven regression models, in which the variables included in the model were decided *a priori*. These included: a linear term for a child’s school grade (to explore the effect of increasing age, where Kindergarten grade is replaced with a zero and grade treated as a numeric term), attendance duration (full-day or half-day, to explore the effect of reduced time spent at school), sex (to identify any sex difference in risk within this age group), and vaccination status (to estimate vaccine efficacy). Random effect terms for school and district were included to identify variation in risk between schools and districts. To assess the robustness of our hypothesis-driven modelling, we also performed an exploration of alternative models using a forward selection process in which the final model’s variables were selecting from a series of candidate variables (described in supplement; Tables S2 and S3).

Data for reproduction of major results and figures is made available through the DRYAD archive [upon publication DRYAD archive number made available].

## RESULTS

### Recruitment

A total of 2,519 students participated in the study at one elementary school (grades K-6), a combined school (grades K-8), and a high school (grades 9-12) from the charter district–District A, and from five elementary schools (grades K-4) and an intermediate school (grades 5-6) in the public school district–District B (**Table 1**). As there were only 12 students in the 12th grade, we combined grades 11 and 12 for all analyses (termed ‘11+’). We achieved good participation rates across the schools: 95.2% in school A1, 99.6% in school A2; 96.9% in school A3; 76.5% in school B1; 89.6% in school B2; 85.0% in school B3; 90.0% in school B4; 84.5% in school B5; 86.8% in school B6. An additional three students dropped out during the study period. After accounting for children for whom we could not achieve follow-up following an absence (see below), we were able to successfully surveil 2,077 children during the study period.

**Table 1:** Demography of the study population

|  | Total | Grade |  |  |  |  |  |  |  |  |  |  |  |
| --- | --- | --- | --- | --- | --- | --- | --- | --- | --- | --- | --- | --- | --- |
|  |  | K | 1 | 2 | 3 | 4 | 5 | 6 | 7 | 8 | 9 | 10 | 11+ |
| **Total study population** | 2519 | 314 | 319 | 343 | 355 | 351 | 227 | 275 | 46 | 38 | 88 | 90 | 79 |
| **Observable for ILI (%)** |  |  |  |  |  |  |  |  |  |  |  |  |  |
| **Yes** | 2077 (82) | 279 (88) | 267 (84) | 300 (87) | 301 (85) | 304 (87) | 185 (81) | 216 (79) | 34 (74) | 30 (79) | 61(69) | 50 (55) | 50 (63) |
| **No** | 442 (18) | 35 (22) | 46 (26) | 43 (23) | 54 (15) | 47 (23) | 42 (19) | 59 (21) | 12 (26) | 8 (21) | 27 (31) | 40 (45) | 29 (37) |
| **Observable population** |  |  |  |  |  |  |  |  |  |  |  |  |  |
| **District** |  |  |  |  |  |  |  |  |  |  |  |  |  |
| **A** | 647 | 54 | 53 | 62 | 56 | 66 | 62 | 69 | 34 | 30 | 61 | 50 | 50 |
| **B** | 1430 | 225 | 214 | 238 | 245 | 238 | 123 | 147 | - | - | - | - | - |
| **School** |  |  |  |  |  |  |  |  |  |  |  |  |  |
| **A1** | 204 | 27 | 26 | 32 | 21 | 32 | 30 | 36 | - | - | - | - | - |
| **A2** | 161 | - | - | - | - | - | - | - | - | - | 61 | 50 | 50 |
| **A3** | 282 | 27 | 27 | 30 | 35 | 34 | 32 | 33 | 34 | 30 | - | - | - |
| **B1** | 201 | 40 | 40 | 36 | 50 | 35 | - | - | - | - | - | - | - |
| **B2** | 270 | - | - | - | - | - | 123 | 147 | - | - | - | - | - |
| **B3** | 240 | 47 | 43 | 54 | 41 | 55 | - | - | - | - | - | - | - |
| **B4** | 188 | 36 | 34 | 44 | 33 | 41 | - | - | - | - | - | - | - |
| **B5** | 342 | 67 | 60 | 59 | 85 | 71 | - | - | - | - | - | - | - |
| **B6** | 189 | 35 | 37 | 45 | 36 | 36 | - | - | - | - | - | - | - |
| **Sex** |  |  |  |  |  |  |  |  |  |  |  |  |  |
| **Female** | 995 | 145 | 125 | 149 | 137 | 136 | 91 | 97 | 14 | 17 | 34 | 27 | 23 |
| **Male** | 1082 | 134 | 142 | 151 | 164 | 168 | 94 | 119 | 20 | 13 | 27 | 23 | 27 |
| **Self-reported vaccination §** |  |  |  |  |  |  |  |  |  |  |  |  |  |
| **No** | 850 | 133 | 111 | 131 | 118 | 122 | 74 | 63 | 17 | 10 | 22 | 25 | 24 |
| **Yes** | 694 | 63 | 95 | 105 | 107 | 120 | 66 | 74 | 10 | 8 | 20 | 13 | 13 |
| **Not reported** | 533 | 83 | 61 | 64 | 76 | 62 | 45 | 79 | 7 | 12 | 19 | 12 | 13 |

§ Of 2077 participants, we obtained a valid immunization response from 1544 children: we failed to interview 296 children regarding their vaccination status; of the
children interviewed, 237 did not know their vaccination status or refused to provide a valid answer for any of the times they were questioned.

### Absenteeism

In total, 1,772 children generated 4,720 absence events during the study period. We were unable to successfully call the home and determine the health status of 442 (24.9%) absent children who did not present to the school nurse with illness; collectively, they were responsible for 970 (20.6%) absence events. Our ability to successfully characterize illness in children following absence was greatest for schools in district A and among younger children (Table S2), We were able to characterize fully 3,750 absence events absences among 1,330 children. Older children (grades 7-12) had fewer absences with ascertained cause (i.e. observable for ILI outcome) (see Table 1).

### Time course of ILI and confirmed infections

Of the children who were absent from school or reported to the school nurses, 408 were diagnosed with ILI and swabbed, of which 271 children were PCR positive for at least one virus. Some children had multiple ILI episodes and were swabbed more than once: 39 were swabbed twice, and six swabbed three times. Of those 271 children testing positive at least once with confirmed virus, 180 (66.4%) were positive for influenza (either subtype A or B), while 56 (20.7%) and 132 (48.7%) were positive for influenza A and B, respectively, during the surveillance period. A small wave of influenza A preceded a larger wave of influenza B cases, corresponding to distinct epidemics of A and B occurring in the wider community (Figure 1). A substantial proportion of children with were positive for respiratory pathogens other than influenza. Among the children testing positive for any virus, 64 (23.6%) were positive for coronavirus, 54 (19.9%) rhinovirus, 28 (10.3%) respiratory syncytial virus (RSV), 6 (2.2%) adenovirus, 3 (1.1%) picornavirus, and 3 (1.1%) metapneumovirus.

**Figure 1.**
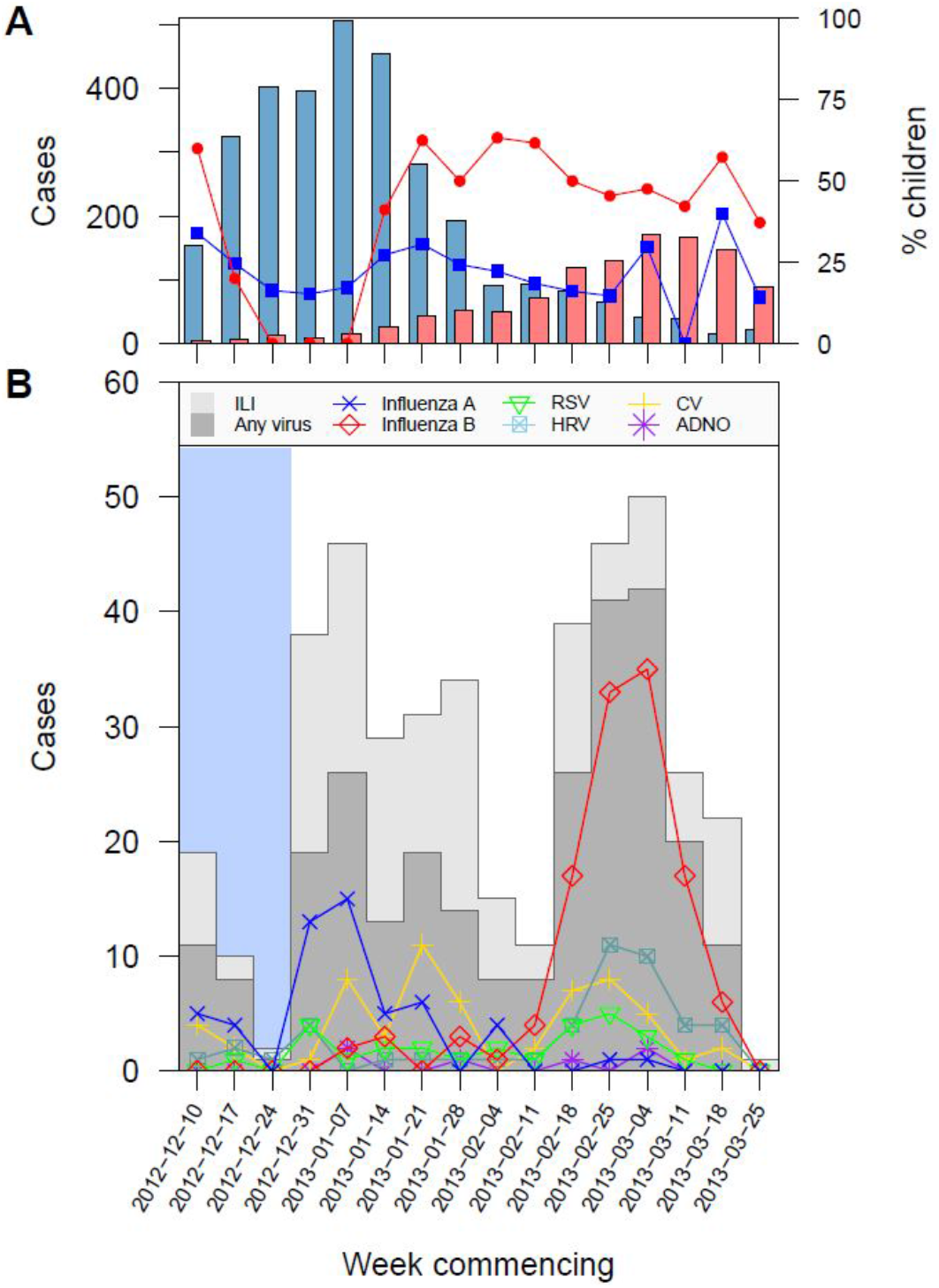
Time series of cases during 2012-2013. (A) Weekly influenza cases reported during the study period by Pennsylvania Health Department for Allegheny county (emergency department and outpatient provider-reported), stratified by influenza type (blue bars, influenza A; red bars, influenza B). Also shown are the percentage of cases who are children (<16 years old); blue and red lines represent influenza A and B respectively. (B) Cases of illness and infection recorded by the study. The pale blue region indicates the period (during 2012) in which only three schools were participating in the study. Numbers of students matching the definition of ILI are denoted by light grey bars; those with virologically confirmed virus are denoted by dark grey bars. Numbers of students testing positive for specific respiratory viruses are shown by the lines and are grouped as follows: influenza A/H1 and influenza A/H3 (Influenza A); influenza B (Influenza B); RSV A and B (RSV); rhinovirus (HRV); coronavirus 229E, NL63, HKU1 and OC43 (CV); adenovirus B, C and E (ADNO).

### Cumulative attack rates (CARs)

In our study, we found an overall cumulative attack rate of 19.7% (95% CI, 18.0−21.5%) for ILI, 13.1% (11.7−14.6%) for infection with a respiratory virus, and 8.7% (7.5−10.0%) for influenza infection. We found significant variation in CAR between grades and schools for children identified with ILI, children testing positive for any of the viruses tested, and children testing positive for influenza (Figure 2; Figures S1, S2, S3). The highest rate of ILI was in grades 1 (26.6%, 95% CI 21.4%-32.3%) and 2 (25.3%, 20.5%-30.7%), while the rate ranged between 10.3% (95% CI, 7.0%-14.4%) and 30.7% (24.2%-37.8%) between schools. Children in grade 1 had the highest CAR for all viruses (20.6%, 95% CI 15.9%-26.0%), and the rate decreased with increasing grade, although kindergarten children had a lower CAR than children in grade 1 (8.2, 95% CI, 5.3%-12.1%). We also found significant differences in CAR for infection by any respiratory virus between the schools (Figure 2B). For influenza, similar patterns were observed: there were significant differences in CAR between grades and between schools, with the highest CAR being in younger aged children in grades 1 to 4, and CAR ranging in individual schools from 3.2% (95% CI, 1.5%-6.0%) to 17.4% (95% CI, 12.4%-23.4%). There were different patterns of CAR for influenza A and B (Figure S1) and other respiratory viruses across grades and schools (Figure S2), and across grade within the same school (Figure S3).

**Figure 2.**
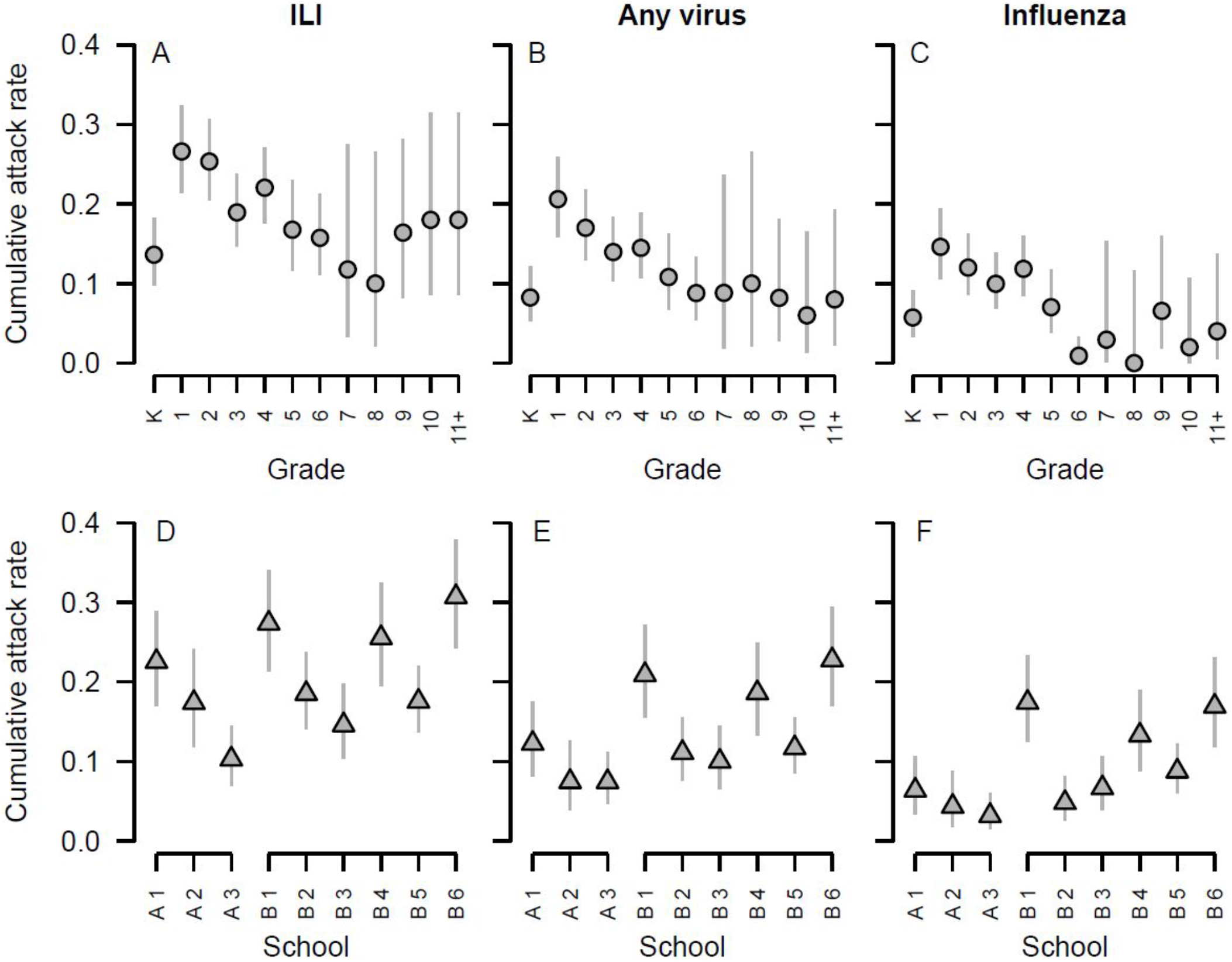
Cumulative attack rate, by grade and school, respectively, for all children diagnosed with influenza-like illness symptoms (ILI) (A, D), those that are laboratory-confirmed with any of the respiratory viruses tested (B, E), and those positive for influenza virus (C, F). Lines denote binomial 95% confidence intervals. Schools are arranged by district (A1 to A3, B1 to B6).

### Sampling delays

Long delays between symptom onset and sampling may introduce bias in the surveillance of acute respiratory viruses, as viral shedding rates diminish as individuals recover. While the delay between symptom onset and swabbing in our study ranged between 0 and 22 days, the median delay was 4 days and 83% of samples were taken within 6 days of symptom onset. Overall, we found no strong indication that positivity rates declined with increase delay in sampling (Figure S3).

### Regression models

CARs are a crude measure of true infection rates, due to the potential for confounders and the pooling of data across all participating schools. To explore the impact of multiple factors in influencing the infection rates of children, we fitted a series of mixed-effect regression models that explicitly incorporated the hierarchical nature of the observations within schools and districts. We tested whether case status (either having ILI, any virus, influenza or specific influenza subtype) was associated with the sex, school grade, self-reported vaccination status of students, or the duration of their instruction at school (full day *vs* half day). Half day duration only occurred in one of the school districts studies (district B).

We found broad agreement in the effect sizes of covariates and random variables across the five modelled outcomes (Table 2). Across all the different models, there was greater variation between schools than between districts. The proportion of variation associated with schools ranged from 11.2% (respiratory virus infection) to 26.5% (influenza B infection). Increasing school grade, a proxy for age, was associated with a reduction in risk in all outcomes. We did a significant effect of sex in any of the models. Vaccination status was not associated with viral infection risk but was associated with a reduction in the risk of ILI. Kindergarten children attending school for a half day were at significantly reduced risk of all infection outcomes except for influenza A infection. Half day attendance was most strongly associated with the risk of influenzas B infection, representing a reduction in risk of between 55% and 92%. These associations were supported further when we used a model selection process to consider alternative model formulations (Tables S2 and S3; supplementary information).

**Table 2.** Odds ratios^$^ of ILI, any virus, influenza (A or B), influenza A, influenza B on length of school instruction, grade, sex, and self-reported vaccination status among schoolchildren in Pittsburgh, PA. Variables significant at the 95% level are shown in bold.

|  | Response variable* |  |  |  |  |
| --- | --- | --- | --- | --- | --- |
|  | Symptomatic infection |  | Laboratory confirmed infection |  |  |
|  | ILI | Any respiratory virus% | Influenza (A or B) | Influenza A | Influenza B |
| **Random effect variables** |  |  | Percent of variance attributable to random effect^ |  |  |
| School | 12.8 | 11.2 | 20.1 | 17.0 | 26.5 |
| District | 0.0 | 1.5 | 1.9 | 0.0 | 9.6 |
| Residuals | 87.2 | 87.3 | 78.0 | 83.0 | 63.9 |
| **Fixed effect variables** |  |  | Odds ratio (95% Confidence Intervals) |  |  |
| Intercept | **0.43 (0.29-0.65)** | **0.28 (0.16-0.49)** | **0.19 (0.09-0.40)** | **0.06 (0.03-0.14)** | **0.10 (0.04-0.25)** |
| Duration of attendance |  |  |  |  |  |
| Full day | 1 | 1 | 1 | 1 | 1 |
| Half day | **0.35 (0.21-0.57)** | **0.28 (0.15-0.51)** | **0.20 (0.10-0.43)** | 0.38 (0.12-1.17) | **0.19 (0.08-0.45)** |
| Grade (linear term) | **0.93 (0.87-1.00)** | **0.91 (0.84-0.99)** | **0.85 (0.77-0.95)** | **0.85 (0.72-0.99)** | **0.88 (0.78-1.00)** |
| Sex |  |  |  |  |  |
| Male | 1 | 1 | 1 | 1 | 1 |
| Female | 0.91 (0.73-1.13) | 0.81 (0.62-1.05) | 0.94 (0.69-1.29) | 1.00 (0.59-1.70) | 0.90 (0.63-1.29) |
| Vaccination |  |  |  |  |  |
| No | 1 | 1 | 1 | 1 | 1 |
| Yes | **0.70 (0.54-0.90)** | 0.80 (0.59-1.08) | 0.81 (0.57-1.17) | 0.59 (0.32-1.12) | 0.90 (0.60-1.36) |
| Not reported | 0.76 (0.58-1.01) | 0.74 (0.53-1.03) | 0.80 (0.54-1.20) | 0.63 (0.32-1.25) | 0.86 (0.54-1.37) |

\$ Odd ratios were calculated using random effect logistic regression
% Influenza A/H1, Influenza A/H3, Influenza B, RSV A, B, Picornavirus 1, 2, 3, 4, Metapneumovirus, Rhinovirus detected, Adenovirus B, C, E, Coronavirus 229E, NL63, HKU1, OC43 detected using Genmark Diagnostic's RVP-RUO panel.
^ Percentage of variance attributable to each random effect was calculated by dividing the standard deviation of each component by the total

### Additional analyses of half-day kindergarten attendance

To further explore the impact of half-day school attendance, we fitted models which explore the association between grade (Table S5) and duration of attendance (Table S6) and infection outcome, in schools with half day kindergarten attendance. We also fitted models of infection outcomes with variables for attendance duration, grade, sex and vaccination status, in schools which have both full day and half day kindergarten students (Table S7). We found Kindergarten students at significantly reduced risk of infection compared to higher (older) grade students in schools with half day attendance (Table S5), though in the same schools, we did not find a significant reduction in risk associated with half day attendance when we did not include in the models (Table S6). However, when we fitted the full model (including terms for attendance, grade, sex and vaccination status) to the four schools in our study with both half and full day kindergarten students, we found half day kindergarten students were at significantly reduced risk of all infection outcomes (Table S7), though we did not find a significant association between infection risk and grade in these models.

## Discussion

We conducted surveillance for influenza-like illness within nine schools during a single influenza season (2012-2013). Nearly 20% of surveilled children experienced at least one episode of ILI during the study period; several experienced multiple episodes. Influenza was the most common viral pathogen identified among these children, although we found the presence of other respiratory viruses in a substantial proportion of detected ILI episodes, suggesting that the burden of ILI in school age children during the normal influenza season may not be entirely due to infection with influenza. ILI and infections were concentrated in younger children, and CARs varied between schools. Modelling of the risk of infection with respiratory viruses and, specifically, influenza identified associations with the grade and the attendance duration of children, where younger children are at higher risk of infection than older children, and kindergarten children attending school for half days were at lower risk than kindergarten children attending for full day instruction.

Contacting parents of absent students to document reasons for absence was a major challenge during the study. However, our rate of follow-up for an absence episode (47.2% of cases) compared favorably to an earlier study in which only 28.6% of absences were verified [21], though we did observe variation in documentation rates by school and grade. The primary problem in contacting parents was inaccurate phone numbers. None-the-less, the pattern of influenza-associated absences confirmed during the study period mirror emergency department and outpatient provider-reported, virologically confirmed influenza from the Allegheny County Department of Health, suggesting that absentee-triggered surveillance was an effective methodology.

Our study found overall CARs for respiratory virus infection of 13.1% (95% CI, 11.7−14.6%) and for influenza infection of 8.7% (95% CI, 7.5%−10.0%); these rates are consistent with those found in other studies of children in non-clinical settings [16]. Our study also shows that the cumulative attack rate of respiratory viruses decreases with increasing grade of school-age children. This age pattern of infection suggests that respiratory infectious disease prevention measures, including efforts to further increase seasonal influenza vaccine uptake, should be especially emphasized for the younger age groups: kindergarten through grade 6. We found the strongest association of grade and time spent in school on the risk of infection with influenza B; this was the outcome with the largest number of cases and, thus, the greatest power to detect significant associations. We found infection outcomes were clustered within schools but varied little between districts, after adjusting for other factors. This observation is consistent with a significant proportion of influenza transmission occurring within schools.

Kindergarten students who attended school for half-day duration were at significantly lower risk of respiratory infection suggests the reduced risk of infection in kindergarten students may be due to reduced time spent within school. Further study is required to determine if these findings can be replicated in other settings, and if half-day school attendance would similarly affect respiratory viral infection in other grades. It is possible that our results reflect the experiences and exposures of kindergarten students within a single district and may not be generalizable to other kindergarten students or other age groups and school grades. Nonetheless, the prospect of a truncated school day as a potential alternative to full school closure, as a control option to reduce the transmission of seasonal and pandemic influenza [4, 22], is intriguing. Pre-emptive, school closures are recommended as a countermeasure during severe influenza pandemics [23], but are associated with significant educational, social and economic impacts [24]. Prior studies of the impact of school closures on respiratory viral transmission have focused on observations around planned and unplanned closures [4, 6, 25], but none have characterized grade specific differences or half-day attendance. Further work is required to assess the efficacy of a half-day attendance regime in non-kindergarten grades and in other school populations on influenza infection and transmission.

There are several important limitations to this study. Our detection of virus relied on identification of an ILI episode within a child; we may, therefore, have underestimated the of incidence of viral infections, particularly if infections can be asymptomatic. While we found little evidence of an effect of vaccination status on infection status, we relied on the children to self-report their vaccination status. This may have introduced bias through misclassification error and masked any true effect vaccines may have had in altering individual infection risk. Our findings may also be biased due to differences in our ability to characterize illness in absent children from different ages and schools; ILI in older children may, therefore, be underestimated in our study. A further limitation is that we have not measured other variables that confound or better explain the observation that half-day kindergarten children are at reduced risk of infection. Kindergarten children in particular schools were reported by the school as attending for either a half or full school day; we had no further information on the time actually spent in school by these individuals.

## Conclusions

Our study found ILI and respiratory infections to be common among school age children during the 2012-2013 influenza season, with the youngest children being at highest risk of infection with influenza. We found no evidence of a difference in infection risk between sexes. We found a shortened school day to be associated with a reduced risk of infection, but this effect was only observed in kindergarteners.

## Data Availability

Data for reproduction of major results and figures is made available through the DRYAD archive.

## Acknowledgments

We would like to thank the children, parents, and staff of our study schools for generously supporting and participating in this study. We are grateful to the virology laboratory of Dr. Charles Rinaldo, Jr. at the University of Pittsburgh and to Arlene Bullotta, UPMC Clinical Virology Lab for processing viral specimens, and to Peter Diggle and Chris Jewell for statistical advice. This work was supported by Cooperative Agreement number [1U01CK00179] from the US Centers for Disease Control and Prevention (CDC, www.cdc.gov). The findings and conclusions in this study are solely the responsibility of the authors and do not necessarily represent the official position of CDC. JMR acknowledges additional support from the Engineering and Physical Sciences Research Council (EP/N014499/1).

## Supplementary material

### Laboratory analysis of samples

Media used to store samples was Remel MicroTestTM M4RT© (Thermo Fisher Scientific, Lenexa, KS) and samples were transported within 72 hours via cooler (maintained at 4° C) to the University of Pittsburgh Medical Center Clinical Virology Laboratory. The lab preserved 200ul of the M4 sample aliquots in 800ul lysis buffer (Biomerieux, Durham NC) and stored in the −80 °C freezer prior to sample extraction with the EasyMag (Biomerieux), a magnetic silicon-based extractor. Samples were processed as a batch.

The nucleic acid or eluates were spiked with Genmark Diagnostic’s (Carlsbad, CA) internal control MS2 prior to testing on the eSensor XT-8 instrument (Luminex, Austin, TX) using the respiratory viral panel or RVP-RUO panel. Tests were performed for both influenza A and B, and included subtyping of H1 and H3 influenza A viruses. Testing was also performed for respiratory syncytial virus (RSV A,B), parainfluenza viruses 1-4 (PIV 1-4), human metapneumovirus (HMPV), human rhinovirus (HRV), adenovirus (ADNO B,C,E), and coronavirus (CV 229E,NKL63, HKU1, OC43).

**Table S1.** Grade-specific contact rates. Values are the mean averages reported by students in a cross-sectional contact diary study during 2011-12 from schools in the same school districts (A and B). Students were asked to complete contact diaries in which they listed all individuals with whom they had a face-to-face conversation or had skin-on-skin contact during a school day. The numbers used here are the subset of those encounters which occurred in school.

| Grade | Encounters |
| --- | --- |
| K | 2.037 |
| 1 | 1.389 |
| 2 | 2.655 |
| 3 | 2.206 |
| 4 | 4.720 |
| 5 | 1.756 |
| 6 | 4.151 |
| 7 | 7.645 |
| 8 | 6.356 |
| 9 | 7.328 |
| 10 | 5.007 |
| 11 | 5.042 |
| 12 | 5.812 |

**Table S2.** Percentage of children reported absent for whom follow-up was unachievable, stratified by school, grade and sex.

| School | % | Grade | % | Sex | % |
| --- | --- | --- | --- | --- | --- |
| District A |  | K | 21.1 | Female | 24.0 |
| A1 | 40.3 | 1 | 19.1 | Male | 25.7 |
| A2 | 46.2 | 2 | 17.4 |  |  |
| A3 | 39.4 | 3 | 21.8 |  |  |
|  |  | 4 | 18.7 |  |  |
| District B |  | 5 | 25.1 |  |  |
| B1 | 19.6 | 6 | 29.4 |  |  |
| B2 | 23.3 | 7 | 42.9 |  |  |
| B3 | 9.2 | 8 | 44.4 |  |  |
| B4 | 7.9 | 9 | 38.6 |  |  |
| B5 | 14.7 | 10 | 56.3 |  |  |
| B6 | 6.2 | 11+ | 43.3 |  |  |

### Model selection

We used a forward selection process to identify covariates that best explained our observed pattern of infection across the participants. The Selection process began from an initial model which only included an intercept term. Candidate variables were systematically considered, and model complexity increased only if deemed significant through a likelihood ratio test.

Candidate variables included: length of school instruction (halfday); school grade as a linear term (grade); sex; vaccination status (vacc); grade-specific social contact rate (Kschool); 1 denotes an intercept term. Social contact mixing rate was collected as part of a parallel study, and is a measure of the average daily number of different people encountered by students in grade with the school setting. This variable was included as a candidate variable as contact rate may be associated with risk of infection. Mixed effect logistic regression models were used throughout, with a nested random effect term of school nested within district. p-values refer to the significance of a likelihood ratio test between that model and the model from the previous step. At each iteration, the model with lowest AIC was selected and progression to the next step was conditional on a likelihood ratio test p-value<0.05. The selection process for each outcome variable is shown in Table S3.

The observed pattern of ILI cases was best explained by a model that contained terms for duration of attendance, self-reported vaccination status, and grade. When the outcome was testing positive for any of the candidate respiratory viruses or testing positive for either influenza type, the selected model contained terms for duration of attendance and grade. For testing positive for influenza A, the selected model only included terms for the number of people encountered within school (a protective association, OR 0.78, 95% CI 0.63-0.97). For testing positive for influenza B, the selected model contained a term for duration of attendance, though we note a model also including a term for grade narrowly missed being selected (Table S4).

**Table S3.** Forward model selection process. Final selected models are highlighted in bold.

| Outcome variable | Step | Model | AIC | ΔAIC | p-value |
| --- | --- | --- | --- | --- | --- |
| influenza-like illness | 1 | 1 | 2042.172 | - | - |
|  | 2 | 1 + halfday | 2029.198 | -12.9744 | 0.000109 |
|  | 3 | 1 + halfday + vacc | 2024.485 | -4.71264 | 0.012826 |
|  | 4 | **1 + halfday + vacc + grade** | **2022.614** | **-1.87149** | **0.049113** |
|  | 5 | 1 + halfday + vacc + grade + sex | 2023.864 | 1.250806 | 0.386732 |
| Any respiratory virus% | 1 | 1 | 1596.555 | - | - |
|  | 2 | 1 + halfday | 1582.741 | -13.814 | 6.99E-05 |
|  | 3 | **1 + halfday + grade** | **1579.05** | **-3.69059** | **0.017056** |
|  | 4 | 1 + halfday + grade + sex | 1578.55 | -0.50063 | 0.113801 |
| Influenza (A or B) | 1 | 1 | 1195.39 | - | - |
|  | 2 | 1 + halfday | 1182.681 | -12.7093 | 0.000125 |
|  | 3 | **1 + halfday + grade** | **1175.316** | **-7.36493** | **0.002212** |
|  | 4 | 1 + halfday + grade + Kschool | 1176.726 | 1.409505 | 0.442228 |
| Influenza A | 1 | 1 | 519.8689 | - | - |
|  | 2 | **1 + Kschool** | **515.9552** | **-3.91364** | **0.015024** |
|  | 3 | 1 + Kschool + halfday | 516.5821 | 0.626834 | 0.241269 |
| Influenza B | 1 | 1 | 942.3886 | - | - |
|  | 2 | **1 + halfday** | **929.5643** | **-12.8243** | **0.000118** |
|  | 3 | 1 + halfday + grade | 927.7762 | -1.78813 | 0.051617 |

% Influenza A/H1, influenza A/H3, influenza B, RSV A, B, picornavirus 1, 2, 3, 4, metapneumovirus, rhinovirus detected, adenovirus B, C, E, coronavirus 229E, NL63, HKU1, OC43 detected using Genmark Diagnostic’s RVP-RUO panel.

**Table S4.**
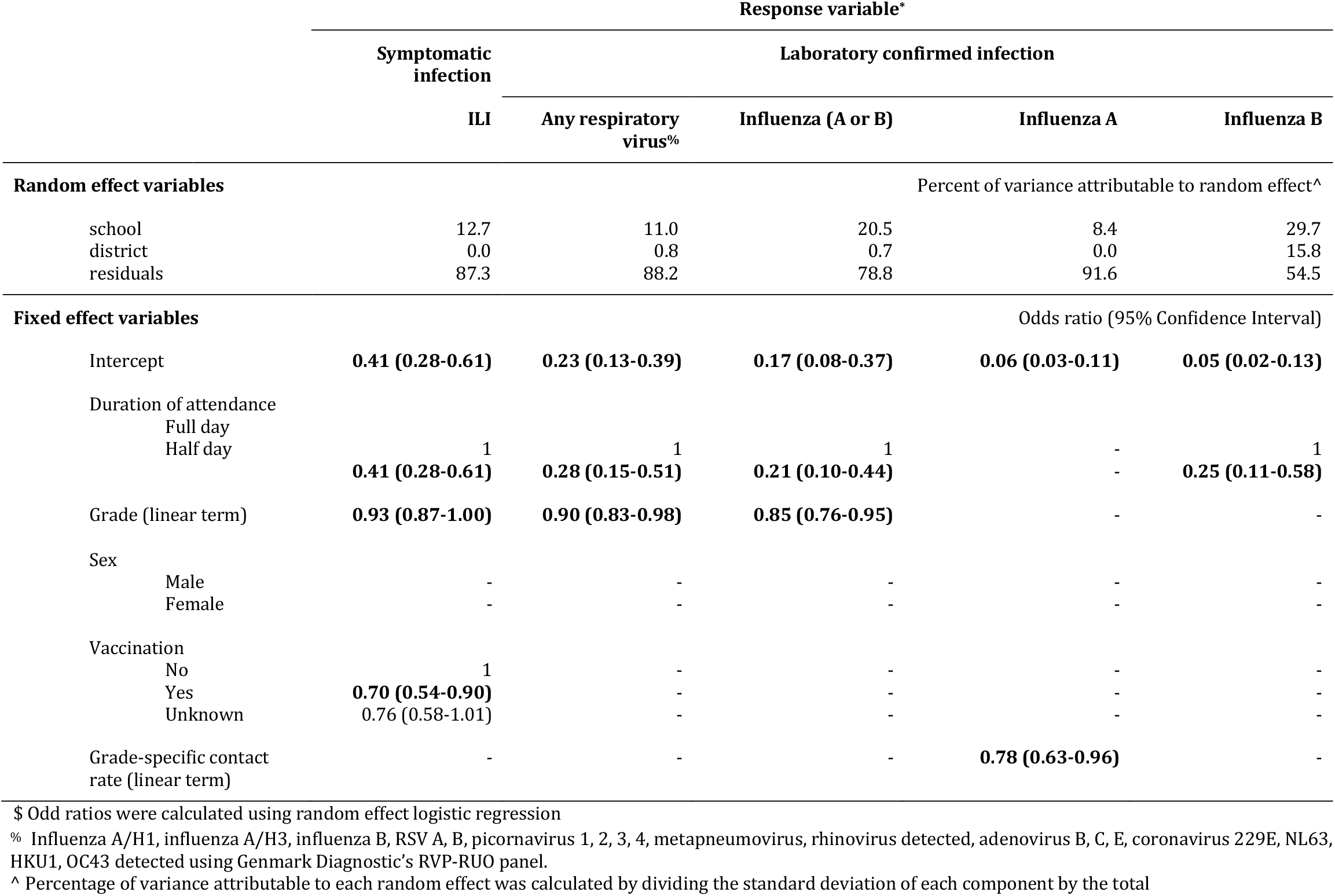
Final models identified using the forward selection process. Hierarchical random intercepts were included for district, and school (within district. Candidate variables included: length of school instruction; grade (linear term); sex; vaccination status; grade-specific social contact rate. Variables significant at the 95% level are shown in bold. Contact rates are the average (mean) number of contacts by grade reported by students.

**Table S5.** Odds ratios^$^ of ILI, any virus, influenza (A or B), influenza A, influenza B on grade within all schools where kindergarteners were taught for half-days (schools B1, B3, B4, B5, and B6).

|  | Response variable |  |  |  |  |
| --- | --- | --- | --- | --- | --- |
|  | Symptomatic infection |  | Laboratory confirmed infection |  |  |
|  | ILI | Any respiratory virus% | Influenza (A or B) | Influenza A | Influenza B |
| **Random effect variables** | Percent of variance attributable to random effect^ |  |  |  |  |
| School | 9.4 | 10.3 | 12.0 | 9.3 | 16.2 |
| Residuals | 90.6 | 89.7 | 88.0 | 90.7 | 83.8 |
| **Fixed effect variables** | Odds ratio (95% Confidence Interval) |  |  |  |  |
| Intercept | **0.14 (0.09-0.23)** | **0.08 (0.05-0.15)** | **0.05 (0.03-0.10)** | **0.02 (0.01-0.06)** | **0.03 (0.01-0.07)** |
| Grade |  |  |  |  |  |
| K | 1 | 1 | 1 | 1 | 1 |
| 1 | **2.61 (1.58-4.31)** | **3.44 (1.90-6.23)** | **3.67 (1.80-7.45)** | 2.61 (0.91-7.52) | **3.91 (1.65-9.27)** |
| 2 | **2.59 (1.58-4.23)** | **2.93 (1.62-5.29)** | **3.26 (1.60-6.61)** | 2.16 (0.74-6.31) | **3.99 (1.70-9.36)** |
| 3 | **1.86 (1.12-3.08)** | **2.19 (1.19-4.02)** | **2.51 (1.22-5.17)** | 0.36 (0.07-1.85) | **3.70 (1.57-8.69)** |
| 4 | **2.17 (1.32-3.58)** | **2.31 (1.26-4.25)** | **3.01 (1.47-6.15)** | 0.76 (0.20-2.86) | **4.14 (1.77-9.70)** |

\$ Odd ratios were calculated using mixed effect logistic regression using a random effect for school, fixed variables are grade.
% Influenza A/H1, influenza A/H3, influenza B, RSV A, B, picornavirus 1, 2, 3, 4, metapneumovirus, rhinovirus detected, adenovirus B, C, E, coronavirus 229E, NL63, HKU1, OC43 detected using Genmark Diagnostic's RVP-RUO panel.

**Table S6.** Odds ratios^$^ of ILI, any virus, influenza (A or B), influenza A, influenza B on duration of instruction within schools with any kindergarteners taught for half-days (schools B1, B3, B4, B5, and B6).

|  | Response variable |  |  |  |  |
| --- | --- | --- | --- | --- | --- |
|  | Symptomatic infection |  | Laboratory confirmed infection |  |  |
|  | ILI | Any respiratory virus% | Influenza (A or B) | Influenza A | Influenza B |
| **Random effect variables** | Percent of variance attributable to random effect^ |  |  |  |  |
| School | 20.8 | 35.6 | 82.1 | 31.3 | 68.1 |
| Residuals | 79.3 | 64.4 | 17.9 | 68.7 | 31.9 |
| **Fixed effect variables** | Odds ratio (95% Confidence Interval) |  |  |  |  |
| Intercept | **0.20 (0.06-0.65)** | **0.16 (0.04-0.65)** | 0.11 (0.01-1.14) | **0.04 (0.00-0.34)** | **0.05 (0.00-0.54)** |
| Duration of attendance |  |  |  |  |  |
| Full day | 1 | 1 | 1 | 1 | 1 |
| Half day | 0.66 (0.20-2.22) | 0.40 (0.09-1.73) | 0.16 (0.02-1.48) | 0.44 (0.03-5.46) | 0.35 (0.03-4.95) |

\$ Odd ratios were calculated using mixed effect logistic regression using a random effect for school, fixed variables are grade.
% Influenza A/H1, influenza A/H3, influenza B, RSV A, B, picornavirus 1, 2, 3, 4, metapneumovirus, rhinovirus detected, adenovirus B, C, E, coronavirus 229E, NL63, HKU1, OC43 detected using Genmark Diagnostic's RVP-RUO panel.

**Table S7.**
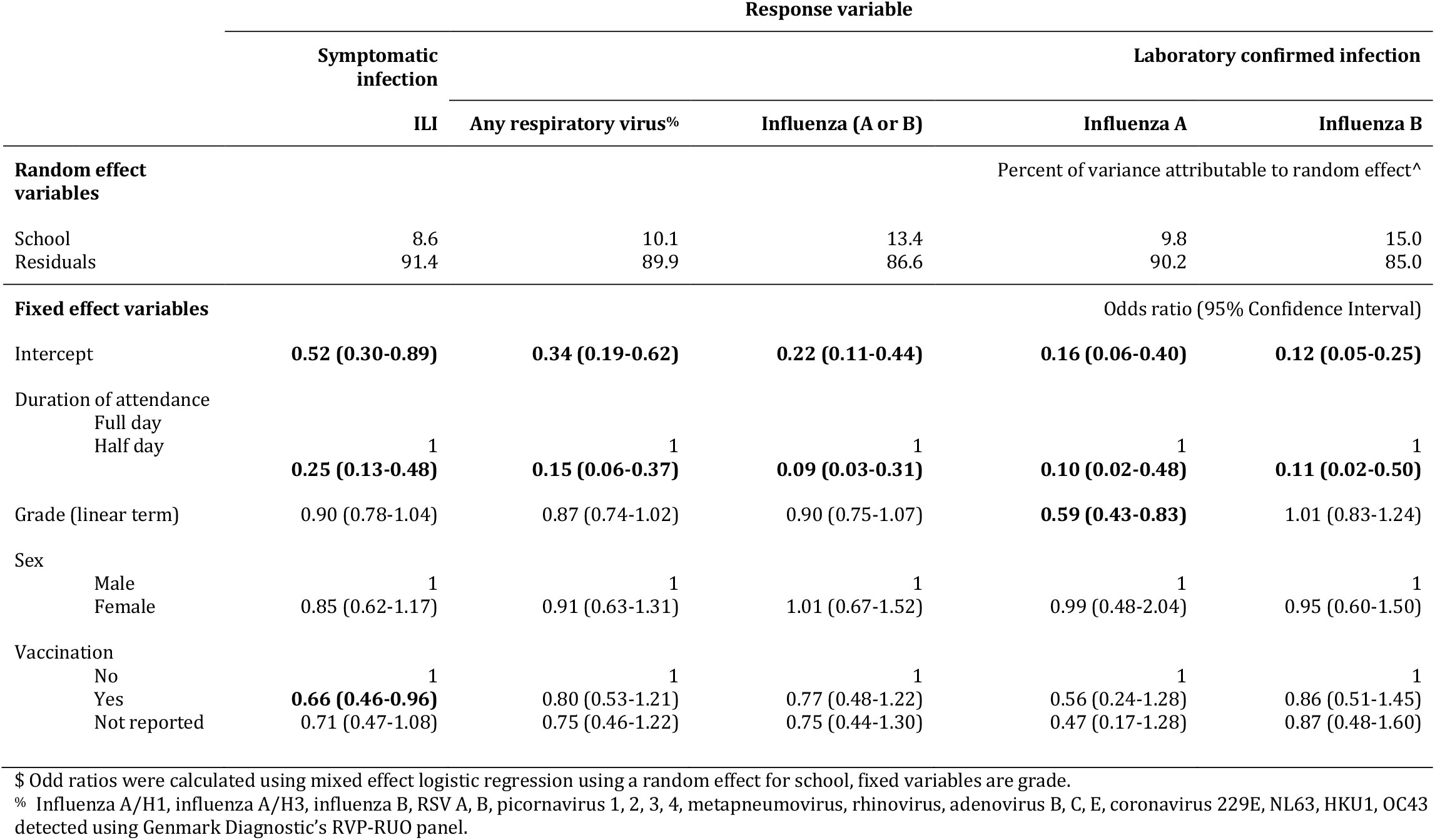
Odds ratios^$^ of ILI, any virus, influenza (A or B), influenza A, influenza B on length of school instruction, grade, sex, and self-reported vaccination status among schoolchildren in schools with both half and full day Kindergarteners (schools B1, B3, B4, and B5). As all the schools belonged to the same school district, only a random effect term for school was included in the models. Variables significant at the 95% level are shown in bold.

**Figure S1.**
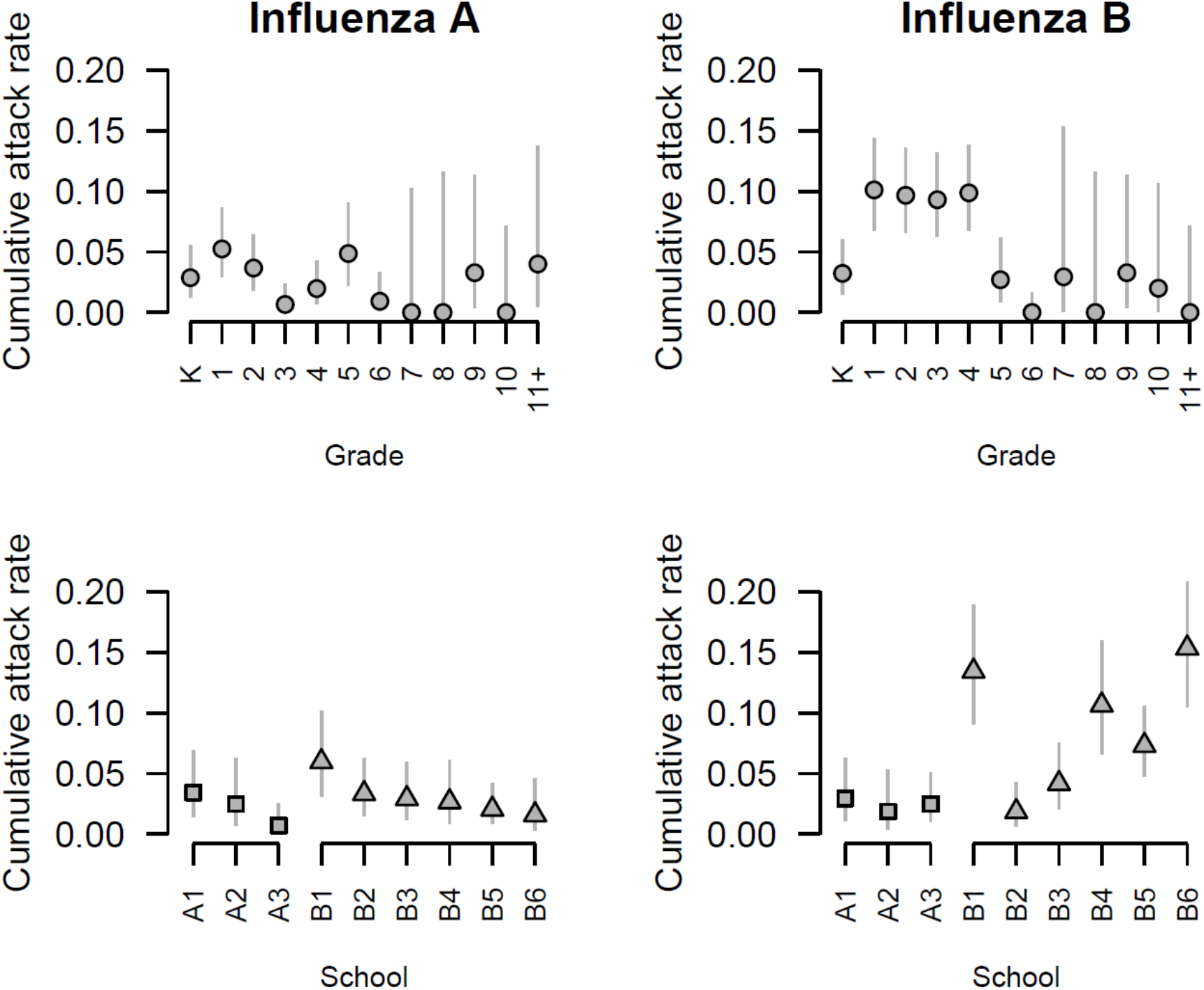
Cumulative attack rate for influenza type A and B, by grade and district. Lines denote binomial 95% confidence intervals.

**Figure S2.**
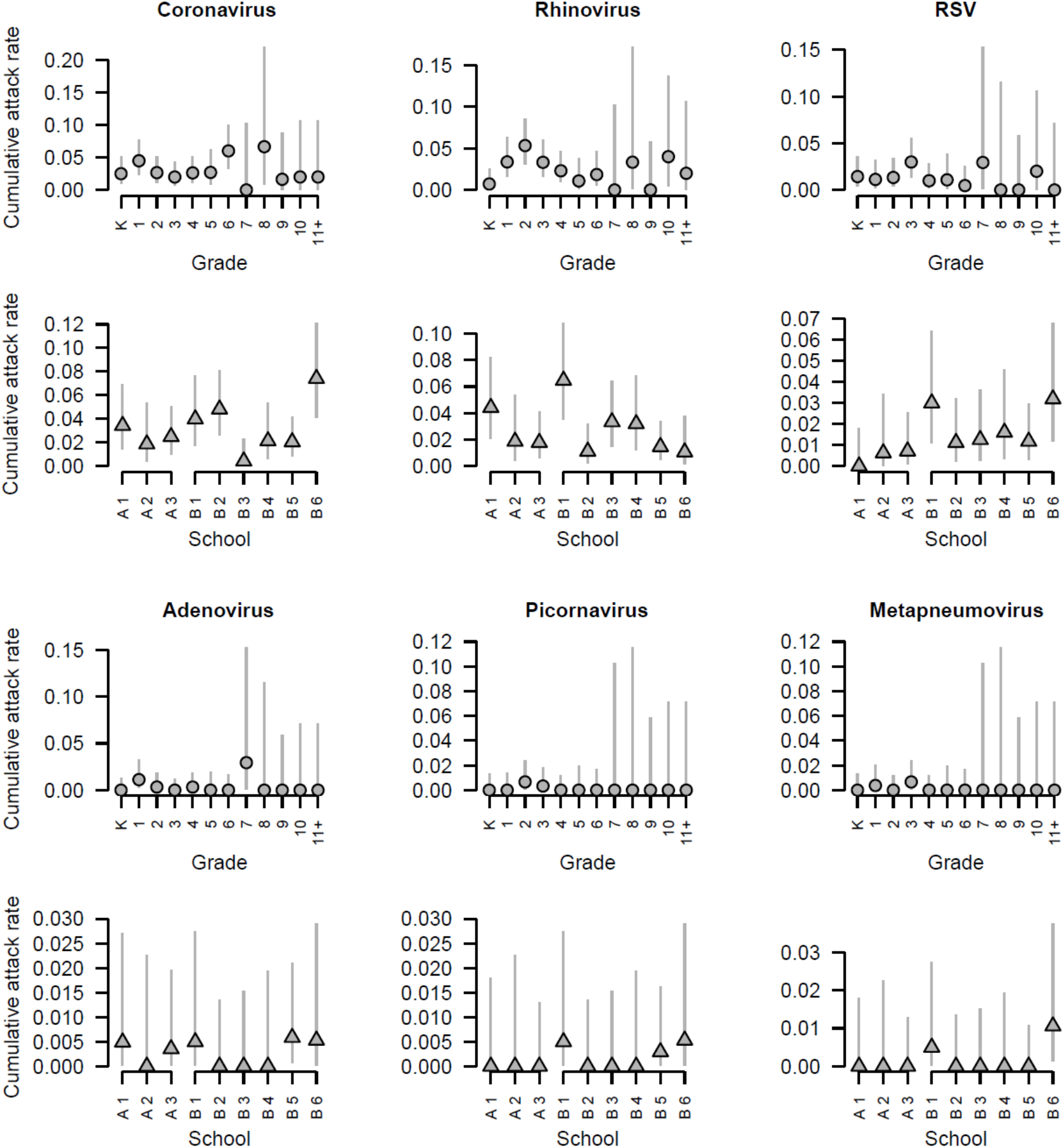
Cumulative attack rates, stratified by grade and school, for other respiratory viruses. Lines denote binomial 95% confidence intervals.

**Figure S3.**
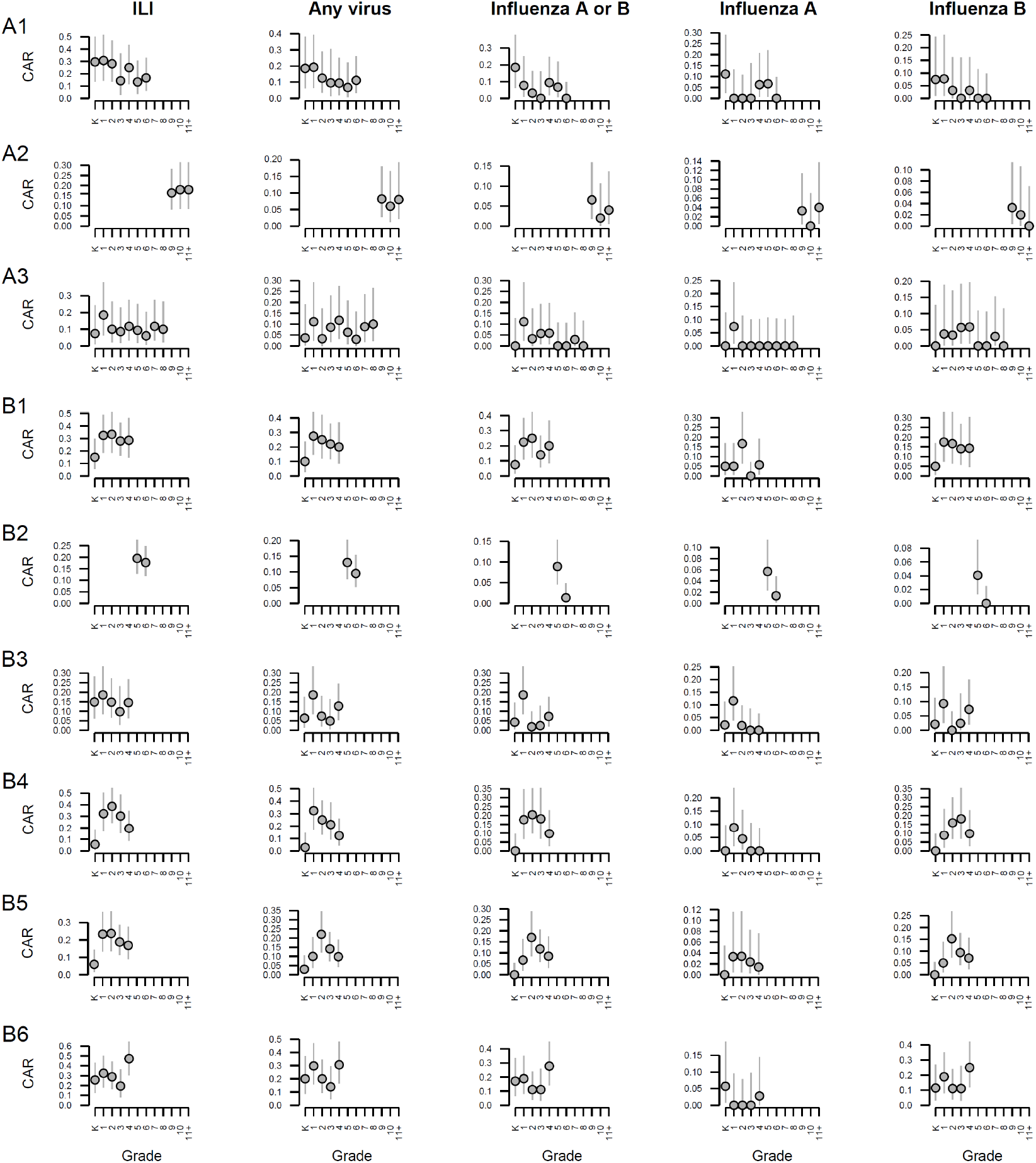
Cumulative attack rate (CAR) by infection outcome (columns), stratified by grade and school (rows).

**Figure S4.**
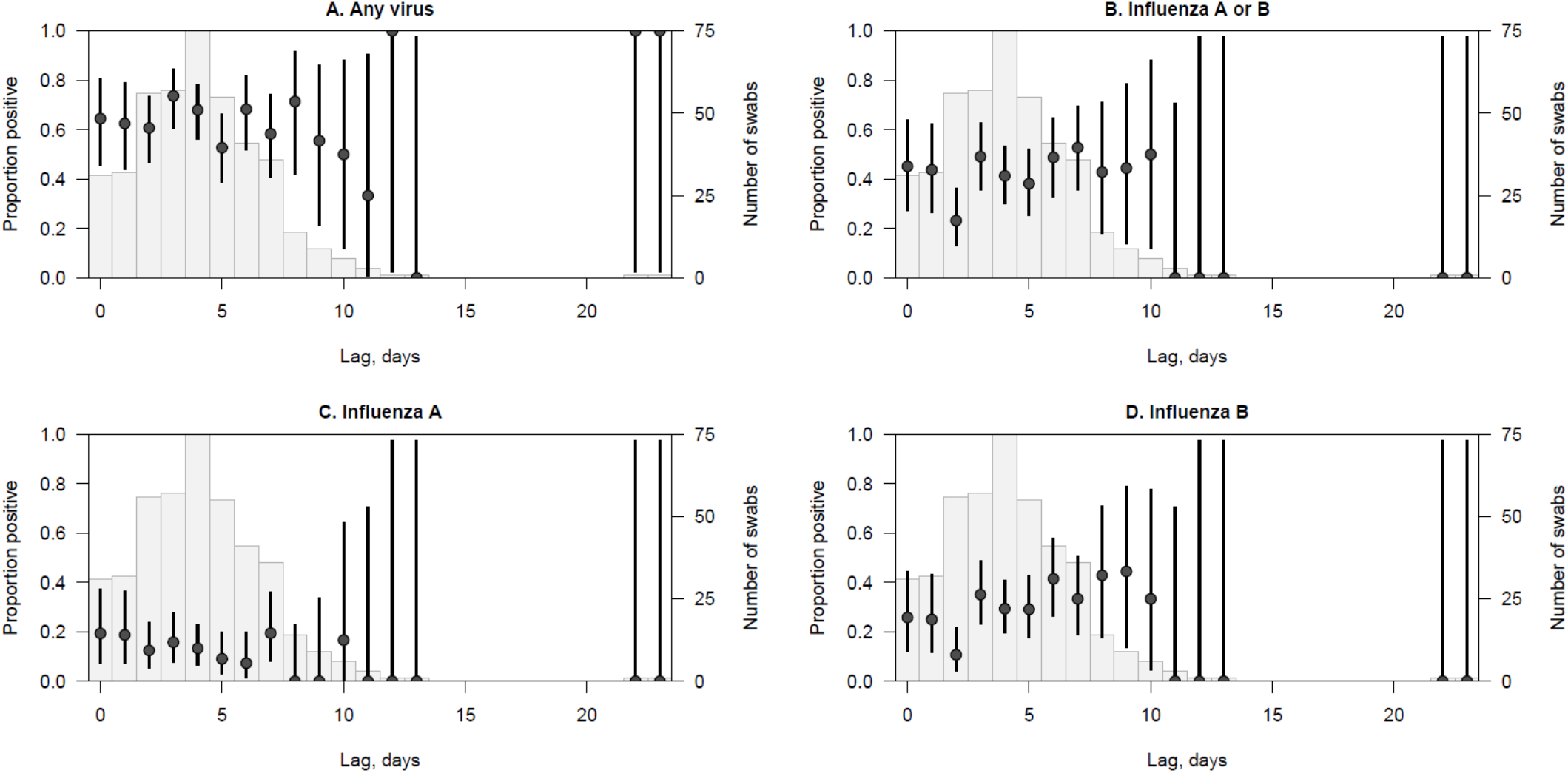
Relationship between the proportion of samples testing positive for virus and the delay between symptom onset date and swabbing date. Circles denote proportions; lines denote 95% binomial confidence intervals. Pale grey bars indicate the number of swabs taken for a given lag.

